# Transcutaneous Spinal Stimulation and Short-burst Interval Treadmill Training in Children with Cerebral Palsy: A Pilot Study

**DOI:** 10.1101/2023.11.21.23298538

**Authors:** Siddhi R. Shrivastav, Charlotte R. DeVol, Victoria M. Landrum, Kristie F. Bjornson, Desiree Roge, Katherine M. Steele, Chet T. Moritz

## Abstract

The purpose of this pilot study was to evaluate the effects of transcutaneous spinal cord stimulation (tSCS) and short-burst interval locomotor treadmill training (SBLTT) on spasticity and mobility in children with cerebral palsy (CP). We employed a single-arm design with two interventions: SBLTT only, and tSCS + SBLTT, in four children with CP. Children received 24-sessions each of SBLTT only and tSCS + SBLTT. Spasticity, neuromuscular coordination, and walking function were evaluated before, immediately after, and 8-weeks following each intervention. Spasticity, measured via the Modified Ashworth Scale, reduced in four lower-extremity muscles after tSCS + SBLTT (1.40 ± 0.22,) more than following SBLTT only (0.43 ± 0.39). One-minute walk test distance was maintained during both interventions. tSCS + SBLTT led to improvements in peak hip and knee peak extension (4.9 ± 7.3° and 6.5 ± 7.7°), that drove increases in joint dynamic range of 4.3 ± 2.4° and 3.8 ± 8.7° at the hip and knee, respectively. Children and parents reported reduction in fatigue and improved gait outcomes after tSCS + SBLTT. Improvements in spasticity and walking function were sustained for 8-weeks of after tSCS + SBLTT. These preliminary results suggest that tSCS + SBLTT may improve spasticity while simultaneously maintaining neuromuscular coordination and walking function in ambulatory children with CP. Significance: This work provides preliminary evidence on the effects of tSCS and the combination of tSCS + SBLTT in children with CP.

## INTRODUCTION

Cerebral palsy (CP) is a disorder of movement and posture caused by non-progressive damage to the developing brain. While CP is primarily a neurological disorder, it also affects the development of neuromuscular and skeletal systems, which negatively impacts mobility and participation in daily activities [1], [2], [3]. Development of corticospinal circuits are impacted in CP, leading to secondary complications such as altered motor control and muscle spasticity [4], [5].

Eighty-five percent of children with CP present with spasticity [6], which is characterized by a velocity-dependent increase in muscle tone. Spasticity is a major contributor to reduced function and increased discomfort in children with CP, limiting gross motor function during activities such as walking [7]. Current spasticity treatments such as baclofen and selective dorsal rhizotomies reduce spasticity but do not consistently translate to improved muscle activity and walking function without extensive additional rehabilitation [8], [9], [10], [11]. Moreover, the long-term effects of these treatments on motor function are rather mixed [12], [13], [14]. Historically, treatments that reduce spasticity have not consistently led to significant improvements in gross motor function [15]. In ambulatory children with cerebral palsy (CP), strength rather than spasticity is more critical for function, with a strong correlation between strength and functional outcomes [16]. Procedures like SDR which sever spinal roots that weaken the connections between muscles and the spinal cord, may explain why walking function often does not improve after SDR unless additional interventions are provided. New interventions that can simultaneously reduce spasticity and improve walking function are needed.

Non-invasive neuromodulation may be an alternative approach that can improve outcomes in CP when combined with physical therapy. Transcutaneous spinal cord stimulation (tSCS) is a novel, non-invasive neuromodulation technique that can modulate spinal and supraspinal circuits [17], [18] especially when implemented with physical therapy [19]. Use of tSCS with physical therapy has reduced spasticity and improved motor function in children and adults with spinal cord injury, multiple sclerosis and CP [19-26]. In children with CP, a single session of tSCS improved coordination of walking and muscle activation [27], while repeated sessions with bodyweight supported treadmill training or activity-based neurorehabilitation therapy improved walking biomechanics and gross motor function, respectively [21], [22], [28].

While these recent results support using tSCS for children with CP, there is limited evidence about the impacts on spasticity, as well as lab- and community-based walking function in ambulatory children with CP. We hypothesized that tSCS may have simultaneous benefits to spasticity and mobility for children with CP. For example, prior work proposes that tSCS may guide reorganization of the spinal and supraspinal circuits by amplifying sensory feedback to support organization of neural pathways [18]. Thus, tSCS may target different mechanisms than current spasticity treatments, such as botulinum toxin type-A injections, baclofen, and selective dorsal rhizotomy. These treatments attempt to reduce muscle activity by inhibiting neural pathways, but often have side effects that result in inconsistent and unsatisfactory changes to walking function without the addition of other interventions, such as physical therapy [29].

The objective of this pilot study was to evaluate the effects of tSCS on spasticity and mobility in children with CP. We evaluated tSCS combined with a unique physical therapy routine specifically designed to improve mobility in children with CP, short-burst interval locomotor treadmill training (SBLTT), compared to SBLTT alone. SBLTT is an experimental treatment for children with CP, which improves walking speed, endurance, and community walking [30]. We hypothesized that tSCS combined with SBLTT would reduce spasticity and improve motor function more than SBLTT alone, and result in increased joint range of motion and reduced demand on muscle activity during walking and community mobility. To test this hypothesis, we compared outcome measures at the completion of each intervention, and at 8-weeks following each intervention.

## METHODS

### Study Design

We conducted a single-arm pilot study with two interventions (Figure 1). All participants received the interventions in the same order with 24 sessions of SBLTT first followed by 24 sessions of tSCS combined with SBLTT (tSCS + SBLTT). Outcomes were collected at the beginning and end of each intervention phase to evaluate the treatment effect of each intervention and at an 8-week follow-up timepoint after each intervention. The 8-week follow-up was chosen because previous literature on physical therapy and spinal stimulation shows that effects last for 6-10 weeks after treatment ends, allowing us to explore the sustained impact of the interventions [22], [30]. The follow-up from SBLTT only (Follow-up 1) and the pre-tSCS + SBLTT time point are the same assessments. Given the small sample size and the known variability between participants with cerebral palsy, we concluded that treating half the children in this small pilot study with stimulation first would create additional confounds due to the documented long-term effects of spinal stimulation [20]. We therefore chose to deliver stimulation as the second intervention. All visits were conducted at the University of Washington, with one exception. Due to family availability, researchers traveled to the home of one participant (P02) for most training visits, using a family treadmill for SBLTT. The participant visited the lab at least once per week for assessments. All study procedures were approved by the University of Washington Human Subjects Division (IRB identifier: STUDY00008896) and the study was registered at ClinicalTrials.gov (NCT04467437).

**Figure 1.**
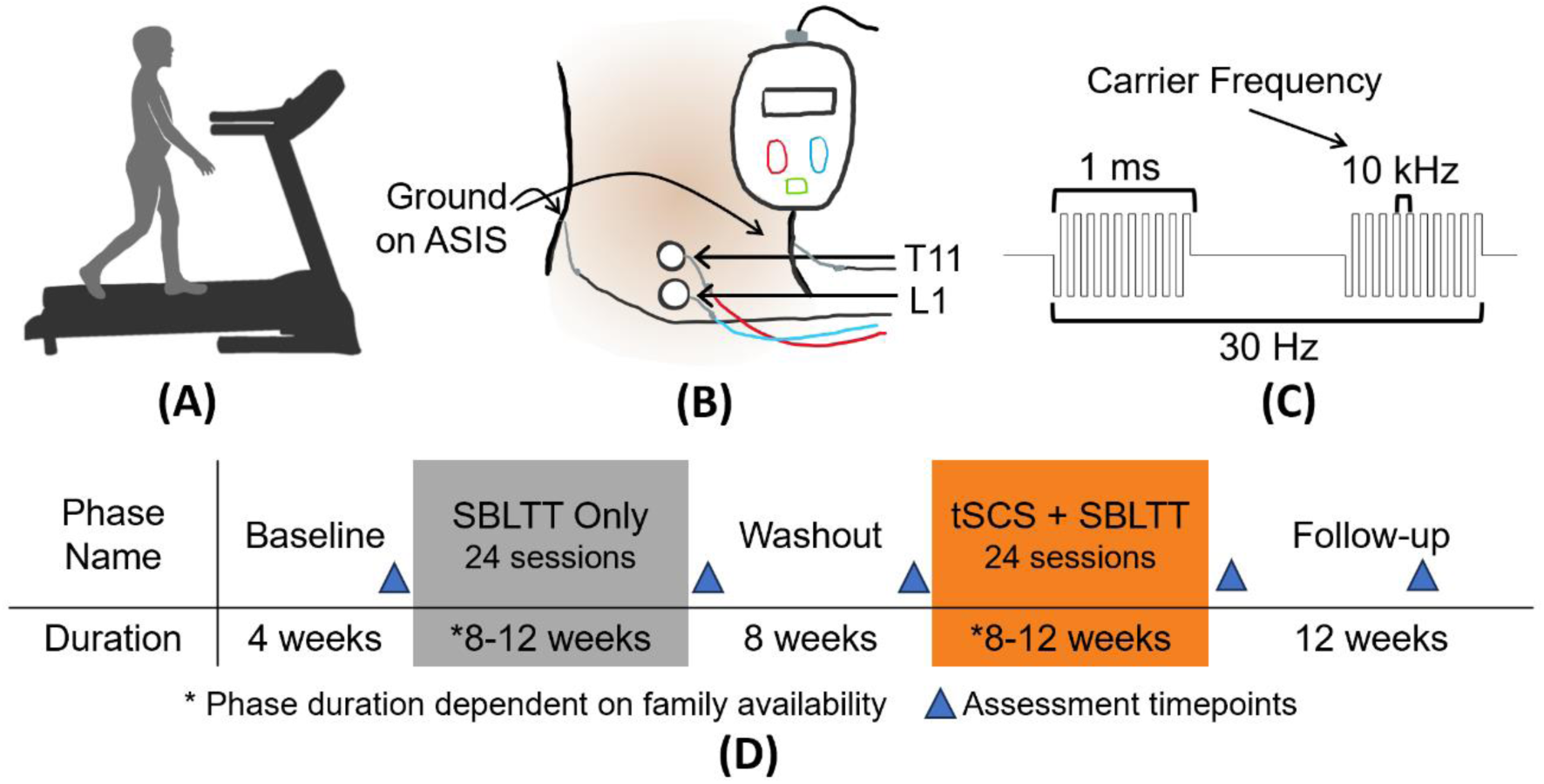
A) Short-burst interval locomotor treadmill training (SBLTT) with contact guard assist. B) Investigative spinal cord neuromodulation device (SpineX, Inc.) with stimulating electrodes on the T11 and L1 dorsal spinous processes and two ground electrodes on the anterior superior iliac spine (ASIS - not visible). C) Spinal stimulation waveform with 10 kHz carrier frequency. D) Protocol timeline including the assessments before and after each intervention and after 8-weeks of follow-up. tSCS = transcutaneous spinal cord stimulation

During both intervention phases, SBLTT was delivered for 30 minutes at each visit following a previously established protocol [19]. SBLTT provides intensive walking practice in which children walk with alternating 30-second bursts of slow and fast speeds, mimicking children’s natural walking patterns. During SBLTT, the slow speed was kept constant across all sessions, while the fast speed was increased within and across sessions based on perceived exertion as measured by both clinical observation and the children’s OMNI Scale of Perceived Exertion [31]. SBLTT was preceded by a 5-to-15-minute active warm-up and concluded with a 5-minute active cool-down. Warm-up and cool-down activities included overground walking, playing, or walking at a low, steady speed on the treadmill. Children wore their orthotic devices that they use in daily life, as described in Table 1, during all interventions for optimal biomechanical alignment during the rigorous SBLTT protocol and to facilitate translation to daily activities. Rest breaks were provided as needed.

**Table 1.** Participant characteristics.

| ID | Sex | Age (years) | Leg Length (m) | Diagnosis | Side | GMFCS | Orthotics | Prior Baclofen (mg)* | SBLTT only |  | tSCS + SBLTT |  | tSCS Amplitude (mA) T11 \| L1 Median (Min, Max) |
| --- | --- | --- | --- | --- | --- | --- | --- | --- | --- | --- | --- | --- | --- |
| | | | | | | | | | Weeks | $\Delta S$ (mph) | Weeks | $\Delta S$ (mph) | |
| P01 | Male | 8-13 | 0.81 | Bi-spastic | Left | II | Bilateral AFO-FCs | 5 as needed | 8 | 1.5 | 11 | 1.1 | 54 (12, 54) \| 40 (15, 40) |
| P02 | Male | 2-7 | 0.50 | Bi-spastic | Left | I | Bilateral AFO-FCs | 10 twice per day | 10 | 0.8 | 10 | 0.9 | 55 (20, 60) \| 45 (15, 47) |
| P03 | Male | 8-13 | 0.75 | Bi-spastic | Right | II | Bilateral AFO-FCs | 10 twice per day | 11 | 1.8 | 11 | 2.2 | 20 (15, 30) \| 25 (20, 30) |
| P04 | Male | 8-13 | 0.85 | Uni-spastic | Left | I | Left lift | None | 10 | 2.3 | 9 | 1.6 | 40 (10, 40) \| 30 (20, 30) |

*The more-affected side is based on the side with more spasticity at baseline and parent reports. ID = participant identifier; Bi-spastic = bilateral spastic CP; Uni-spastic = unilateral spastic CP; Side = more-affected side; GMFCS = Gross Motor Function Classification System; AFO-FC = ankle foot orthosis footwear combination; $\Delta S$ = Change in fast burst SBLTT speed from first to last session, based on the fastest speed at the first and last session. tSCS amplitude applied to T11 = thoracic spinous process 11, L1 = lumbar spinous process 1. \*Baclofen use was gradually weened to zero under the guidance of the child's primary care physician ending two weeks prior to enrollment in the study, except P01 who took 5 mg of baclofen once during the SBLTT only phase of the study.*

During tSCS + SBLTT, the parameters and application of the investigative spinal cord neuromodulation device (SpineX, Inc.) followed previously reported protocols [27]. We delivered pulses of 1 ms at a frequency of 30 Hz with a 10 kHz carrier frequency. Stimulation was applied using adhesive gel electrodes with the cathodes delivering stimulation placed just below the T11 and L1 spinous processes using 3.2 cm round electrodes to target myotomes of the lower extremity muscles and for consistency with prior work [28]. The anodes, serving as the ground electrodes, were 5.1 x 8.6 cm rectangular electrodes placed bilaterally over the anterior, superior iliac spines (ASIS) (Figure 1B). During each visit, stimulation was applied throughout all activities, including warm-up, SBLTT, rest breaks, and cool-down for an average of 56 ± 10 minutes. Amplitude for the sub-motor threshold stimulation was determined based on three factors for each subject: 1) participant reported sensation beneath the cathodes, 2) children’s self-report of the ease of walking, and 3) a physical therapist’s clinical observation of gait quality and participant’s behavior. The stimulation amplitudes used for each child are reported in Table 1.

### Participants

We enrolled ambulatory children with spastic CP and Gross Motor Function Classification System (GMFCS) Levels I-II who were not currently taking spasticity management medications, did not have a history of selective dorsal rhizotomy, and had not undergone a lower extremity surgery or botulinum toxin injections in the past 1 year. Four children with CP participated in the study (Table I). Two participants, P02 and P03, weaned off their daily use of baclofen 2-weeks before starting the study. Another participant, P01 took baclofen as needed prior to the study and took 5 mg once during the SBLTT phase. Participants had not received any botulinum toxin injections or orthopedic surgery before joining the study, except P01 who had botulinum toxin injections 9 years prior. Children and parents were informed of the study procedures and signed an informed consent and age-appropriate assent form.

### Outcome Measures

Lower limb spasticity and walking distance were the focus of this preliminary study. Outcomes included the Modified Ashworth Scale (MAS), Tardieu Scale, and the 1-minute walk test (1-MWT). MAS was assessed on the hamstrings, quadriceps, gastrocnemius, and soleus muscles bilaterally. MAS scores were converted into an ordinal scale, such that a value of zero indicated no spasticity and a value of five indicated joint rigidity. We also assessed the Tardieu Scale for the hip extensors, knee flexors and extensors, and ankle flexors and extensors as an additional measure of spasticity. Both the MAS and Tardieu scores were averaged across all muscles at each timepoint, with the same assessor each time. Walking capacity in the lab was measured using the 1-minute walk test (1-MWT), which measures the distance walked in one-minute and is considered a reliable measure of functional ability and walking endurance in ambulatory children with CP [32], [33]. One participant, P01 performed a 6-minute walk test at every time point, before we switched to the 1-MWT to enable work with younger participants (P02-4). The 6-MWT distances for P01 were converted to a 1-minute walking distance by calculating the average distance walked in 1-minute for each assessment. These tests were conducted by a Physical Therapist who was not blinded to the study design and interventions.

Biomechanical changes in walking were also assessed at the hip, knee, and ankle joints on each participant’s more-affected side. Joint kinematics and muscle activity were quantified during walking on a 10-meter walkway. Participants were instructed to walk at a self-selected pace while barefoot for a minimum of 25 steps at each assessment timepoint. Lower extremity motion data were collected using a modified Helen-Hayes marker set [23] and a 10- or 12-camera motion capture system at 120 Hz (Qualisys AB, Gothenburg, SE). Data were processed using custom MATLAB scripts (MathWorks, Natick, MA), USA) and OpenSim v4.3 (Stanford, USA) using a 23 degree-of-freedom model scaled to each individual participant [24], [25]. Across trials, the root-mean-square (RMS) and maximum model error for all markers were below 2 cm and 4 cm, respectively, which align with best practices for model quality [26]. Each joint’s dynamic range was calculated as the average change in joint angle across gait cycles.

Electromyography (EMG) data (Delsys Inc, Natick, MA) were synchronously recorded during motion capture trials bilaterally for five muscles: rectus femoris (RF), vastus medialis (VM), biceps femoris (BF), tibialis anterior (TA), and medial gastrocnemius (MG). Using custom MATLAB scripts, raw EMG signals were high pass filtered (4th order Butterworth; 20 Hz), zero-centered, rectified, and low pass-filtered (4th order Butterworth; 10 Hz). Signals were then normalized to the 95th percentile of maximum activation across trials for that day and reported as milli-volts/millivolts (mV/mV). Integrated muscle activity was defined as the area under the curve for predefined phases of the gait cycle [39] and calculated for the VM and MG muscles during stance where muscle activity is usually increased for gait patterns seen in children with CP [40]. Co-contraction of antagonistic muscle pairs was defined as the co-contraction index (CCI) calculated as:

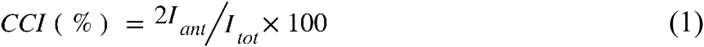

Where *Iant* is the antagonistic muscle activity and *Itot* is the sum of agonist and antagonist EMG activity [41]. The antagonist muscle is the muscle that has lower muscle activity over the duration of a CCI calculation. A lower CCI indicates less co-contraction and more coordinated timing of muscle activation.

Walking capacity and performance were evaluated by lab- and community-based measures. Walking speed (10-meter walk test, 10-MWT), functional mobility (Timed Up and Go, TUG), and dynamic balance (Pediatric Balance Scale, PBS) were evaluated in a lab-setting [42], [43], [44]. Patient-Reported Outcomes Measurement Information System (PROMIS^®^) Pediatric Profile-Fatigue short form was used for child-reported level of fatigue for all participants except P02 whose parent completed the parent proxy form due to the child’s young age [45].

To quantify community-based walking activity, participants wore a step counter (StepWatch, Modus Health, Edmonds, WA) on their left ankle for seven consecutive days. Data from four weekdays and one weekend day were included for the final analyses. Average daily stride rates were calculated for each participant because evidence shows that high stride rates in natural environments indicate higher participation [46]. To evaluate child and parent perceptions about gait outcomes, we used child and parent-reported questionnaires, the Gait Outcomes Assessment List (GOAL) [47]. Total scores were calculated for each participant and parent. Means and standard deviations were calculated for all outcomes. Hatchfill2 was used in MATLAB to provide color patterns in some figures [48].

## RESULTS

We found that tSCS + SBLTT reduced spasticity with a maintenance of walking function in all participants. Spasticity, measured by MAS, improved by 1.4 ± 0.22 after tSCS + SBLTT compared to 0.43 ± 0.39 after SBLTT only. Average MAS scores remained low for 8-weeks after tSCS + SBLTT but not after SBLTT only compared to pre-intervention scores (Figure 2A). Reduced spasticity following tSCS + SBLTT was also indicated by the Tardieu scale. Spasticity reduced by 4.3 ± 3.0 points after SBLTT only and 7.3 ± 4.3 points after tSCS + SBLTT. Reduction in average Tardieu scores nearly sustained through 8-weeks follow-up after tSCS + SBLTT (Figure 2B). The reductions in spasticity of individual muscles were greater across the tSCS + SBLTT intervention compared to SBLTT only except P03, who had greater reductions in hamstrings and quadriceps muscles during SBLTT only (Fig 3). In general, the greatest reductions in spasticity were observed in gastrocnemius and soleus muscles. These muscles both contribute to ankle plantarflexion, with the gastrocnemius muscle also contributing to knee flexion. Reductions in spasticity were also observed at the hamstrings and quadriceps but were more variable across interventions.

**Figure 2.**
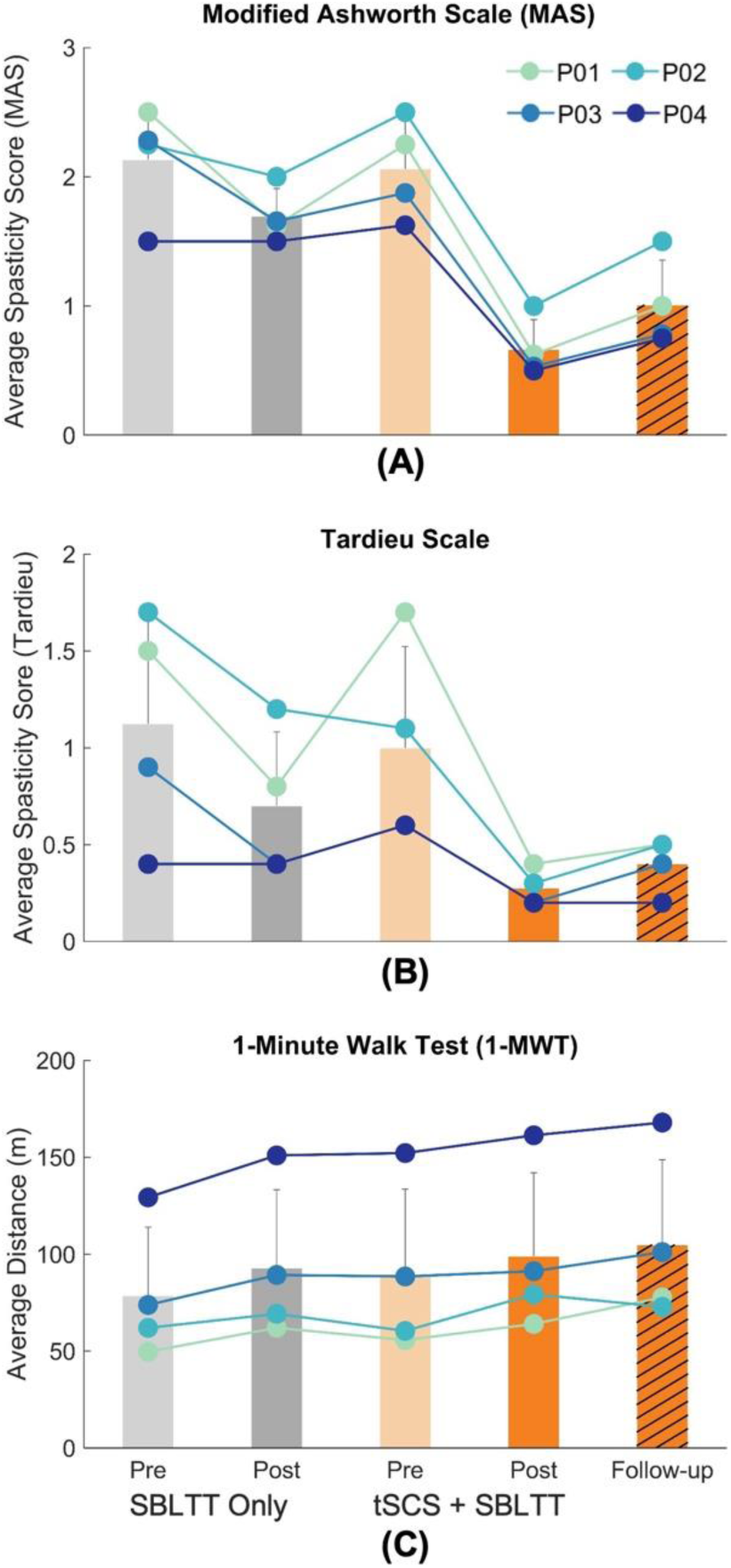
Spasticity outcomes of A) Average Modified Ashworth Scale (MAS) for four muscles bilaterally, B) Tardieu Scale, as well as C) walking distance during the 1-minute walk test (1-MWT) for each participant before and after each intervention and after 8-weeks follow-up.

**Figure 3.**
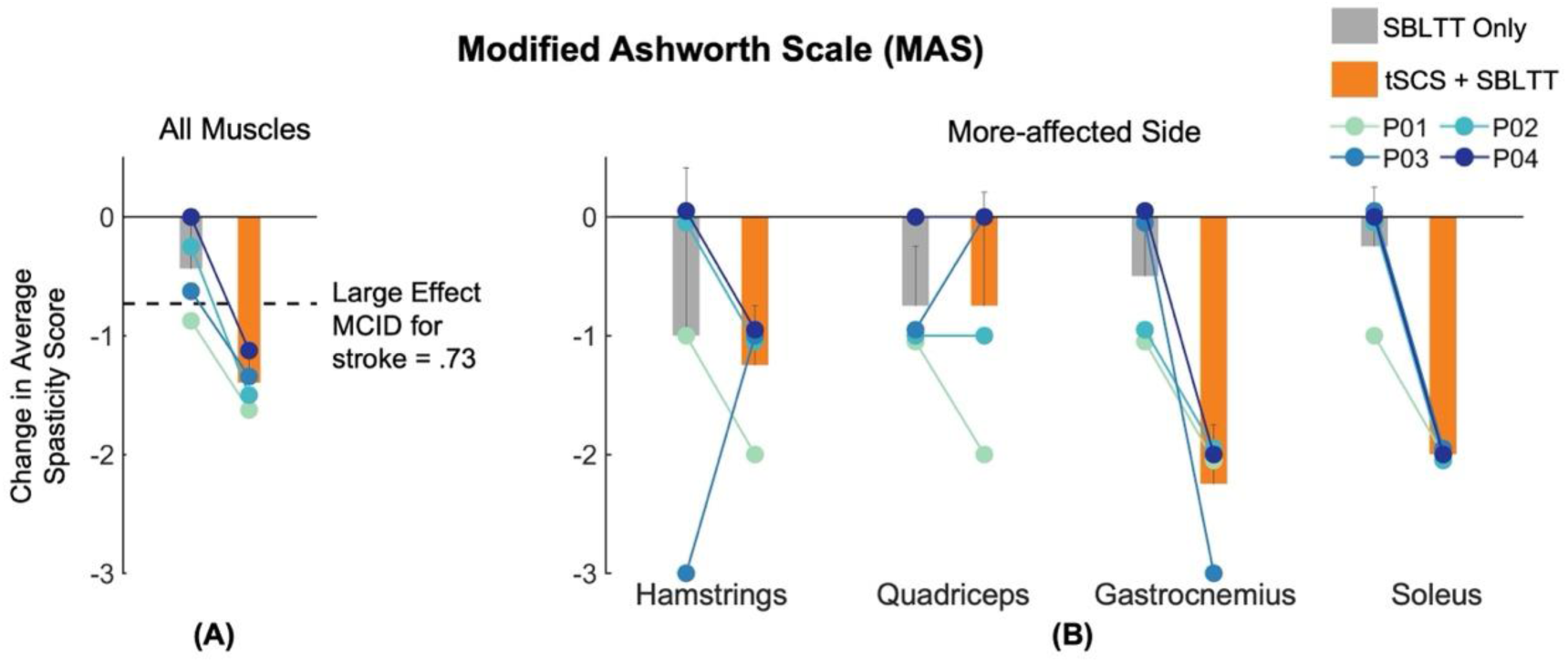
Change in muscle spasticity measured using the Modified Ashworth Scale (MAS). A) Results averaged across all muscles bilaterally, were lower after tSCS + SBLTT than SBLTT only. The horizontal dashed line indicates the minimum clinically important difference (MCID) for spasticity reduction in adults post-stroke [50], as similar values are not available for children with CP. B) Secondary outcomes of individual muscle MAS for the more-affected side of each participant, including the hamstrings, quadriceps, gastrocnemius, and soleus. Larger negative numbers indicate greater reductions, or improvements, in muscle spasticity.

Distance walked during the 1-MWT was maintained throughout both interventions (Figure 2C). We observed a small increase of 14 ± 6 meters (m) after SBLTT only and continued increase of 10 ± 7 m after tSCS + SBLTT. These changes exceeded the large effect size MCID for both GMFCS level I (9 m) and II (8.3m) [49].

We also documented more joint extension and dynamic range of motion during walking after tSCS + SBLTT compared to after SBLTT only (Figure 4). At baseline, participants exhibited increased hip and knee flexion during stance phase of walking, characteristic of crouch gait. During SBLTT only, participants had minimal change in hip and knee peak extension by an average of -0.42 ± 7.3° and 2.4 ± 6.6°, respectively. This may have contributed to a change in the overall joint dynamic range of motion during walking after SBLTT only of 2.0 ± 3.5° and -2.4 ± 11.0° for the hip and knee, respectively. In contrast, after tSCS + SBLTT, participants increased peak joint extension during gait, with average improvements of 4.9 ± 7.3° and 6.5 ± 7.7°, driving increases in joint dynamic range of motion of 4.3 ± 2.4° and 3.8 ± 8.7° at the hip and knee, respectively (Figure 4A and B). Minimal changes were observed in joint kinematics at the ankle (Supplemental Figure 2).

**Figure 4.**
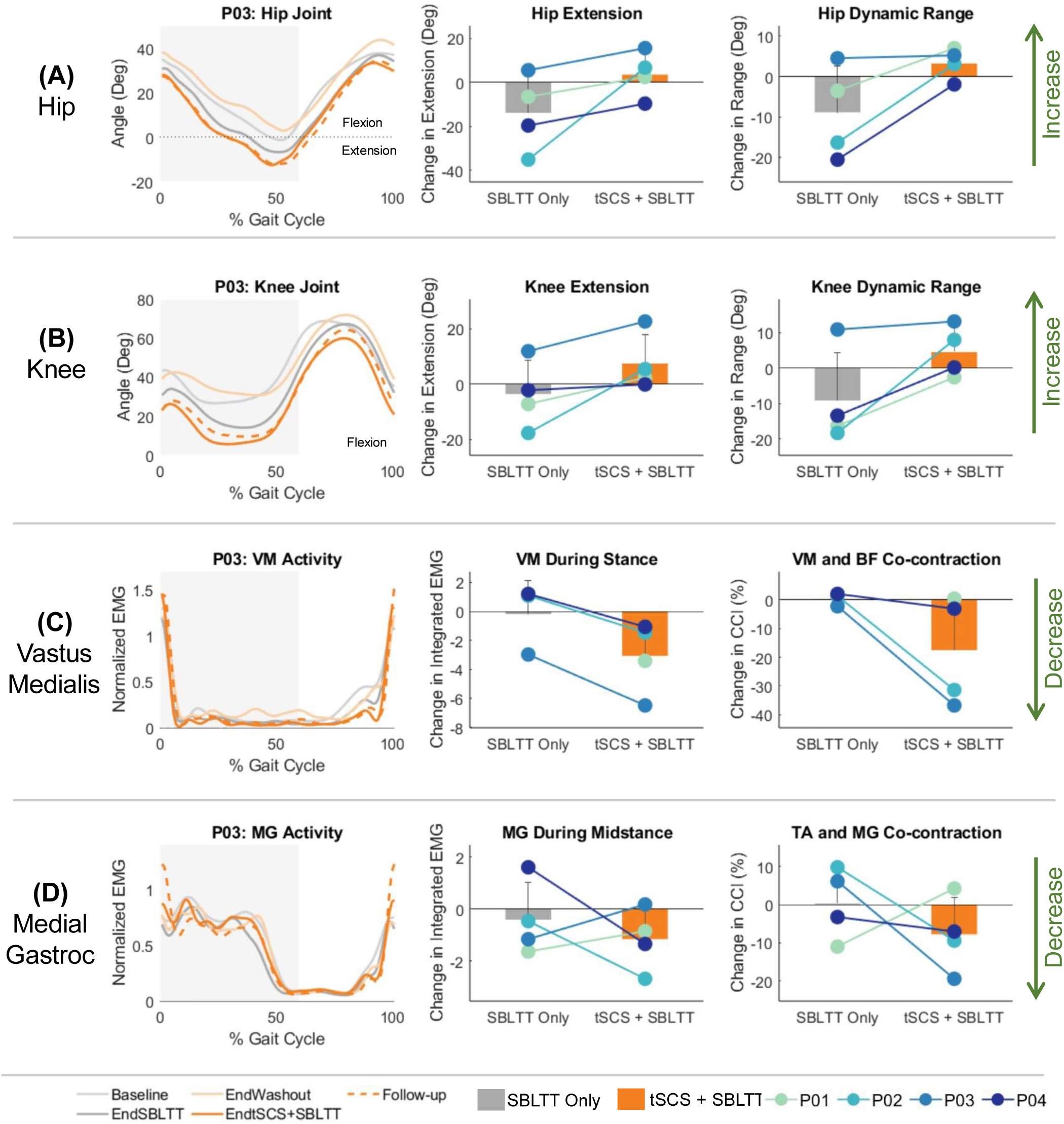
Average joint kinematics and muscle activity during barefoot walking at a self-selected speed. The left column shows examples from P03, other columns represent data from all participants as follows. Arrows on far right indicate the direction of change for middle and right columns. Changes in A) hip and B) knee joint kinematics, C) vastus medialis (VM) activity and co-contraction index (CCI) between the VM and biceps femoris (BF), and D) medial gastrocnemius (MG) activity and CCI between the MG and tibialis anterior (TA). Arrows indicate desired direction of change for each variable, including increased hip/knee extension, increased dynamic range of motion, decreased muscle activity during stance, and decreased co-contraction. Notes: The VM data from P01’s baseline visit is missing from (C) due to poor EMG signal during data collection. The sample VM activity from P03 in (C) also has a consistent large artifact at heel strike, likely due to sensor movement.

In addition to changes in joint mechanics during tSCS + SBLTT, all participants maintained or reduced muscle activity in the VM and MG muscles after tSCS + SBLTT. Change in VM activity for P01 during SBLTT Only was not included in analysis due to a poor EMG signal at the baseline visit. Integrated VM activity during stance increased on average 5.9 ± 12 during SBLTT only and decreased 3.1 ± 2.5 during tSCS + SBLTT. Integrated MG midstance decreased 0.41 ± 1.4 and 1.2 ± 1.2 during SBLTT and tSCS + SBLTT, respectively. This resulted in less co-contraction during tSCS + SBLTT compared to SBLTT only between the VM and BF (SBLTT only: 5.6 ± 11%; tSCS + SBLTT: -18 ± 19%) and between the MG and TA (SBLTT only: 0.43 ± 9.4%; tSCS + SBLTT: -7.9 ± 9.7%).

Participants’ walking speed improved during both interventions. Specifically, walking speed, measured via the 10-MWT improved by 0.06 ± 0.10 meters/second (m/s) after SBLTT only and by 0.16 ± 0.25 m/s after tSCS + SBLTT (Figure 5A). We measured an increase in community mobility following SBLTT + tSCS. Peak stride rate in the community did not change after SBLTT only but improved by 3.0 ± 4.1 strides/minute after tSCS + SBLTT (Figure 5B). Functional mobility and balance were also assessed in the laboratory. Average time taken to complete TUG reduced by 1.3 ± 1.6 seconds after SBLTT only, and further reduced by 0.4 ± 1.0 seconds after tSCS + SBLTT (Supplemental Figure 2B). Improved TUG times after SBLTT exceeded the medium effect size for both GMFCS level (1.1s) and level II (0.7s) [49]. Dynamic balance as evaluated by Pediatric Balance Scale (PBS) scores improved by 3.7 ± 3.2 points after SBLTT only with continued improvements of 3.7 ± 5.5 points after tSCS + SBLTT (Supplemental Figure 2C).

**Figure 5.**
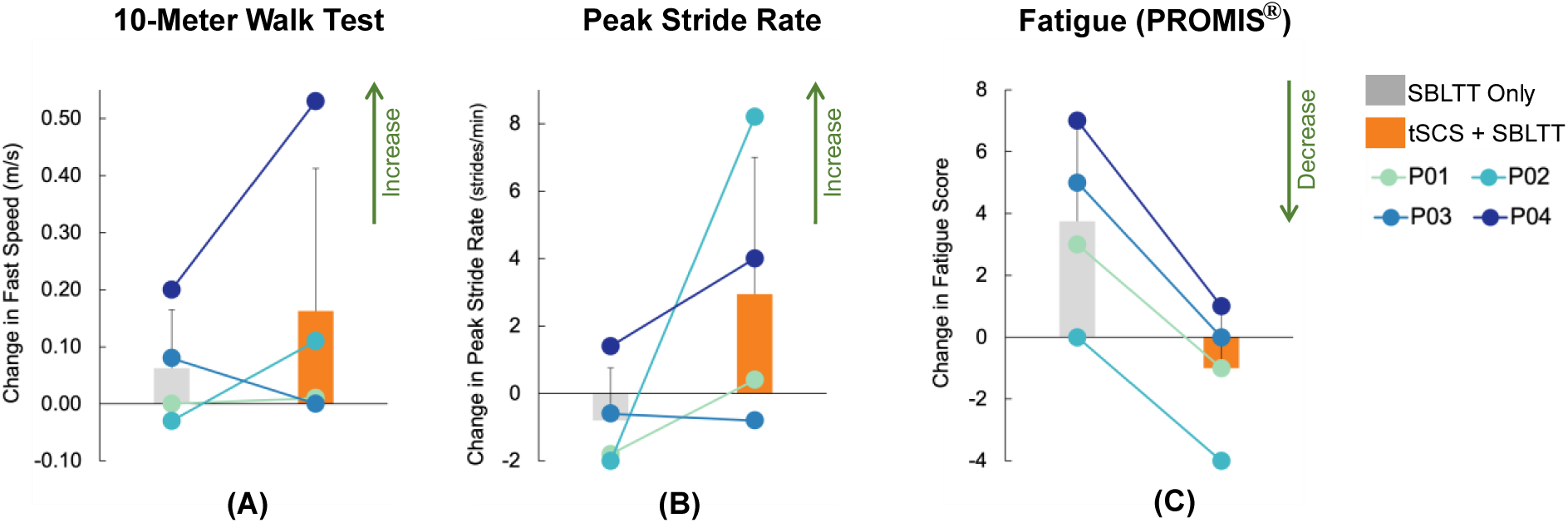
Improvements in walking capacity and performance as measured by A) lab-based walking speed via the 10-meter walk test (10MWT), B) community peak stride rate captured via StepWatch device in children’s natural environments, and C) reduction in self-reported fatigue scores captured via the Patient-Reported Outcomes Measurement Information System (PROMIS®) Pediatric Profile Fatigue short form. Arrows indicate the desired direction for improved function.

All participants reported greater reductions in fatigue after tSCS + SBLTT compared to after SBLTT only. Participants reported a 3.8 ± 3.0 point increase in fatigue after SBLTT only, but a 1.0 ± 2.2 point decrease in fatigue after tSCS + SBLTT as captured via the PROMIS® (Figure 5C). Child-reported gait outcomes scores reduced by 4.3 ± 5.5 points after SBLTT only and increased by 9.7 ± 8.5 points after tSCS + SBLTT. Parent-reported gait outcomes scores reduced by 1.8 ± 0.96 points after SBLTT only but improved by 3.0 ± 4.1 points after tSCS + SBLTT (Figure 6).

**Figure 6.**
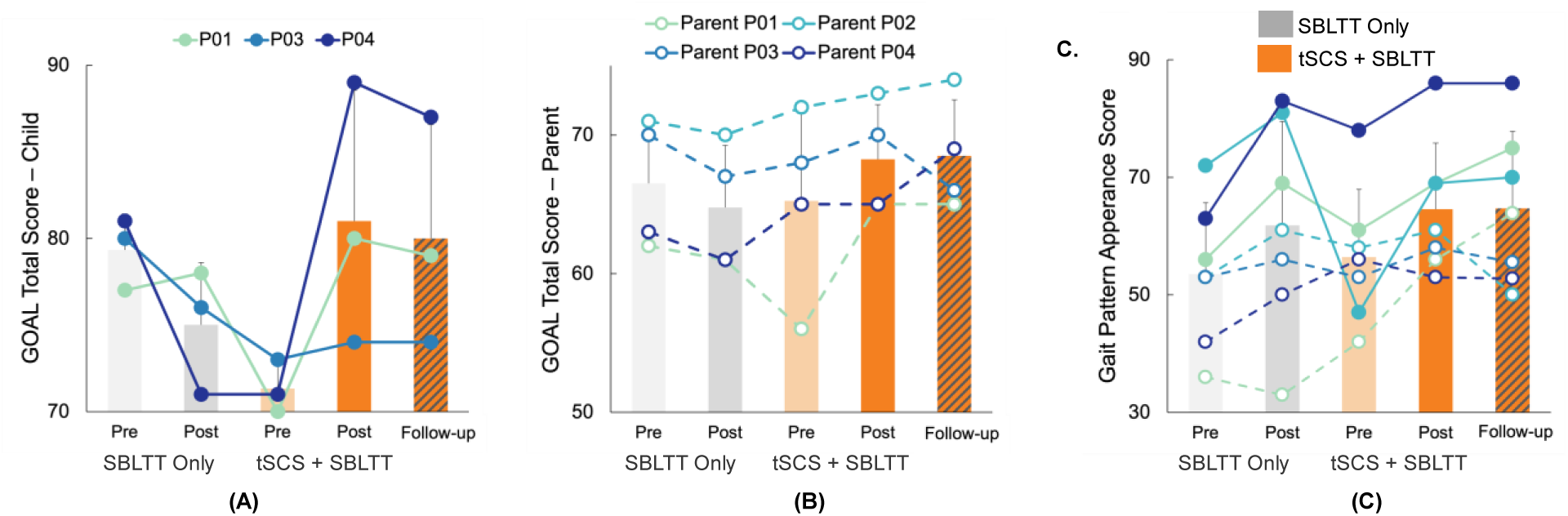
A) Child and B) parent-reported total Gait Outcomes Assessment List (GOAL) scores, and C) Child and parent reported Gait Pattern Appearance domain scores. Higher scores indicate better self-reported gait function. Please note that the y-axis does not begin at zero for all of these GOAL results.

Lastly, there were no serious adverse events during the study. The only minor expected event was mild erythema around cathodes that resolved on its own within 15 minutes after stopping the stimulation, which occurred three times for P01.

## DISCUSSION

The combination of transcutaneous spinal cord stimulation and SBLTT (tSCS + SBLTT) led to greater improvements in average spasticity compared to SBLTT only. tSCS + SBLTT also showed changes in joint dynamic range and changes in muscle activity during walking compared to SBLTT only.

It is important to place the observed changes in spasticity in a clinical context. MAS scores reduced after tSCS + SBLTT more than SBLTT alone. There is no reported minimum clinically important difference (MCID) for the MAS in children with CP. In adults who had a stroke, however, an average change in lower extremity MAS of 0.73 is considered a large effect MCID [50]. All four children with CP in our study achieved this MCID for reduction in spasticity after tSCS + SBLTT, which was also sustained for at least two months with no further study treatment (Figure 3).

It is also interesting to note that families anecdotally reported changes in clinical recommendations following participation in the study. For P02, their physician no longer recommended selective dorsal rhizotomy (SDR) and P03 was told they could remain off spasticity medication for at least another 6 months before reassessment after spasticity reductions observed during the tSCS + SBLTT portion of the study.

Common, clinically available treatments of spasticity for children with CP include botulinum toxin type-A (BTA) injections, baclofen, and SDR surgery. BTA injections are applied intramuscularly and reduce muscle activity by blocking acetylcholine release from motor neurons at the neuromuscular junction [51]. BTA injections provide temporary reduction in spasticity, requiring repeated injections that come with negative effects on muscle development and reduced spasticity response on repeated use [10], [51]. Baclofen is a pharmacological intervention option that can be taken orally or delivered via an implanted intrathecal pump. Baclofen reduces the release of excitatory neurotransmitters in the spinal cord that contribute to spasticity [52].

While these treatments reduce spasticity, they have certain limitations that may constrain their use in children. For example, baclofen may result in epilepsy, anxiety, and sleep disorders [53], [54]. SDR is a neurosurgical procedure that permanently transects afferent nerves in the spinal cord after which children require intensive rehabilitation to recover to pre-SDR function. Most importantly, these spasticity treatments do not have long-term benefits on motor function and spasticity [10], [12], [13]. Our pilot study showed improvements in spasticity with sustained walking function following tSCS + SBLTT with no serious adverse effects. More research with a longer follow-up period is needed to understand if these effects persist.

Our findings built on prior studies of tSCS showing improved function for children with CP where reports on spasticity have been limited [21], [22], [27], [28]. The two prior studies that evaluated spasticity did not report what muscles were assessed and one did not report the change in MAS value [22], [28]. A reduction in spasticity has been observed for four children at GMFCS Level III-V, but no effect of tSCS on spasticity was reported for the two GMFCS Level I-II children included in the study when tSCS was combined with activity-based neurorehabilitation therapy [22] or when tSCS was combined with bodyweight supported treadmill training across GMFCS Levels I-IV [28]. The difference in results could be due to the different physical therapy interventions utilized and how tSCS was applied in the prior studies. One study applied tSCS at lower amplitudes (12-18 mA at C5-6 and 10-16 mA at T11-12) while the other had a wide range of stimulation amplitudes (10-50 mA). In this study, stimulation amplitudes were adjusted based on children’s tolerance and observation of their walking patterns. The type of activity used with tSCS, as well as the stimulation location and amplitude, may influence the efficacy of tSCS in modulating spasticity. This emphasizes the need for future inquiry, with subsequent work required to optimize physical therapy paired with tSCS to positively impact spasticity, mobility, and daily activities.

We also observed greater improvements in hip and knee extension after tSCS + SBLTT compared to SBLTT only. Increases in hip and knee extension are characteristics of reductions in crouch gait [55]. Reductions in spasticity at the hamstrings and gastrocnemius may have driven increased knee extension over 24 sessions of tSCS + SBLTT, but further investigation is needed [56]. These findings are similar to prior work that found improved hip, knee [27], [28], and trunk kinematics [23] in children with CP and SCI.

We also quantified reductions in excessive muscle activity in the vastus medialis during tSCS + SBLTT compared to SBLTT only. Walking with less crouch gait can reduce demand on hip and knee extensors (i.e. integrated area of the VM and MG muscles), as we observed here with less hamstring activity, and potentially reduce fatigue [40]. This could explain why participants reported greater self-reported fatigue after SBLTT only, but not after tSCS + SBLTT. It is also possible that tSCS + SBLTT manifested a placebo effect on self-reported fatigue. Fatigue in CP is associated with deteriorated walking, especially as children transition into adulthood [57]. Therefore, approaches that reduce fatigue are a high priority for the CP community [58].

Interestingly, minimal changes were observed in ankle joint kinematics, despite large improvements in spasticity at the ankle (Supplemental Figure 2). This may be because all participants wore their community assistive devices during SBLTT, including three participants who wore their rigid ankle foot orthoses-footwear combination (AFO-FC) that limits movement and sensation of the ankle. We chose to use the children’s prescribed AFO-FC during SBLTT to maximize transfer to daily activities. The AFO-FCs likely supported the fast-walking speeds achieved during SBLTT but may have also reduced the sensory feedback that tSCS aims to boost during training, and potentially limiting therapeutic effects at the ankle. Evaluating the effects of orthoses on training responses represents an important area for future research.

We also observed some individual improvements in lab-based, community-based and self-reported measures of walking function. MCIDs for the 1-MWT and TUG have been established by Hassani et al. [49]. All participants increased walking distance during the 1-MWT after SBLTT only and reached at least a medium effect of the MCID, with two participants reaching the large effect MCID. All participants also increased their walking distance after tSCS + SBLTT, compared to pre-tSCS + SBLTT, with only two participants reaching the large effect MCID [49]. The variability of changes in walking distance emphasizes that different children with CP may respond differently to tSCS and SBLTT. All participants improved their TUG performance time after SBLTT only, with one participant reaching a large effect MCID and another reaching a medium effect MCID. Three of four participants further improved TUG performance time after tSCS + SBLTT, with two participants reaching the large effect MCID, as reported by Hassani et al. [49]. This suggests that both SBLTT only and tSCS + SBLTT improved walking performance in a clinically important way and tSCS + SBLTT maintained it while spasticity was simultaneously reduced. Moreover, only tSCS + SBLTT led to sustained improvements in spasticity and maintained walking function 8 weeks after the intervention was complete.

Children with CP often have reduced levels of physical activity in daily life and demonstrate less walking intensity compared to typically developing peers [59]. Our early findings suggest that tSCS + SBLTT may facilitate community walking intensity, as shown by higher peak stride rates, which is similar to what has been reported in the SBLTT literature [30]. Improvements in lab-based measures of walking function provide preliminary evidence on the effects of tSCS + SBLTT on walking capacity in a controlled environment, while improvements in peak stride rate captured in the community via a StepWatch provide insight into transference to children’s day-to-day natural environments.

Positive self-reported changes in gait outcomes captured via the GOAL questionnaires provide a holistic view of participants’ and their parents’ positive subjective gait-related experiences after tSCS + SBLTT. Both children and parents reported either an increase or a maintenance in achieving walking goals after tSCS + SBLTT. This was driven by improvements in Domain E: Gait Pattern and Appearance. Self-reported fatigue increased after SBLTT Only but reduced after tSCS + SBLTT. The cause of elevated energy expenditure that contributes to increased fatigue in CP remains unclear and can be due to numerous, interacting factors including changes in motor control, kinematics, and muscle activity [60]. Self-reported measures of fatigue can include perceptions of walking and other activities of daily living. Since tSCS + SBLTT improved hip and knee extension and reduced muscle activity, these changes may have contributed to the observed reductions in reported fatigue, although future work should examine the mechanisms by which tSCS and SBLTT may decrease fatigue for children with CP. Future work should also explore the use of tSCS + SBLTT to understand its implementations and user’s perceptions on how they may affect community mobility.

It is also important to establish the underlying neuromechanical mechanisms driving changes for evidence-driven, personalized rehabilitation. By modulating sensorimotor activity, tSCS aims to induce neuroplasticity, or promote a more natural organization of neural pathways, thereby improving sensory integration and motor control [17], [61]. In children with CP whose early brain injury affects both the spinal and supraspinal circuits [4], [62] disorganization between the supraspinal and the spinal pathways causes inadequate sensorimotor processing [62]. Sensation is often altered, and can have a negative influence on motor function [63], [64]. Impaired sensation further leads to a disruption of inhibitory and excitatory inputs, manifesting as spasticity [65] and impacting mobility [66]. Prior work theorized that the combination of tSCS and motor training promotes reorganization of the spinal-supraspinal connectivity by amplifying sensory signals at the level of the spinal cord during functional activities [18], [22]. Motor practice during this amplified state of sensory feedback may result in improved sensorimotor integration at both spinal and supraspinal levels that is maintained for at least several months following treatment [18]. Our findings provide preliminary support for this hypothesis, allowing for simultaneous improvements in how sensory information is integrated both involuntarily (i.e., spasticity) and voluntarily (i.e., walking). Confirming t he underlying neurophysiological effects of neuromodulation represents an exciting avenue for future work.

Despite encouraging findings, there are several limitations to this work. First, the small sample size and variability in ages of four males with spastic CP and GMFCS I-II limits the generalization of results. The lack of randomization of the intervention arms may have also led to an additive effect of SBLTT across interventions. Given that tSCS is known to have persistent effects that can last for many months after treatment the order of treatment was kept consistent to prevent introducing more cofounding factors into a small sample size [20]. Regardless, larger studies are needed using a randomized treatment order to definitively distinguish between the effects of SBLTT Only and tSCS + SBLTT. A second limitation of this study is that the timing over which each intervention was delivered differed slightly due to family availability and the COVID-19 pandemic. One participant also completed the majority of SBLTT sessions in an integrated home program with two researchers coming to the home. Nonetheless, the number of therapy sessions was the same between all participants and intervention phases and demonstrates the ability of the intervention to adapt to family needs with the aim of reducing burden on families. Third, we did not restrict the physical therapy that participants may have been receiving outside the study or activities during the follow-up period. However, this indicates that even when the interventions are applied in the real -world context of changes to daily life, tSCS + SBLTT consistently resulted in greater improvement in spasticity compared to SBLTT only. Further, we did not measure motor threshold for each participant and thus are unable to report the level of sub-motor threshold stimulation as a percentage of motor threshold. Future work should consider incorporating quantitative methods of measuring motor threshold, such as with electromyography, as an additional method of calibrating the amplitude of stimulation. We also acknowledge that there is a potential for assessor bias, as there was no blinding in the study. Participants were also not blinded to the treatment arms, which may have introduced response bias into more subjective assessments such as GOAL and PROMIS®. Future studies should consider blinding the assessors, especially for subjective measures such as the MAS, and blinding participants to the treatment arms by utilizing sham stimulation. Lastly, the true effect size of tSCS + SBLTT could not be estimated because SBLTT alone was always delivered first.

## CONCLUSION

In this pilot study, we report that the combination of transcutaneous spinal cord stimulation and short-burst interval locomotor treadmill training led to sustained reductions in spasticity for at least 8 weeks in four children with cerebral palsy. During both short-burst interval locomotor treadmill training alone and its combination with stimulation, walking function was maintained despite reductions in spasticity. Children walked with less crouch mechanics, while also reporting improved gait outcomes and reduced fatigue after training with spinal stimulation. Future research should investigate the applicability of these findings to other forms of therapy for children with CP and elucidate the underlying neuromechanics driving improvements. Further studies are needed to quantify how spinal stimulation and physical therapy interventions can be integrated to address the needs and goals of children with CP.

## Data Availability

All data produced in the present study are available upon reasonable request to the authors.

## ACKNOWLEDGEMENT

The authors thank the children and their families for the time they dedicated to the research. We also thank Dr. Soshi Samejima, Rich Henderson, and Lauren Bachman for assisting with interventions and assessments, and Avocet Nagle-Christensen for data analysis.

