## Supplemental Figures for "Transcutaneous Spinal Stimulation and Short-burst Interval Treadmill Training in Children with Cerebral Palsy: A Pilot Study"

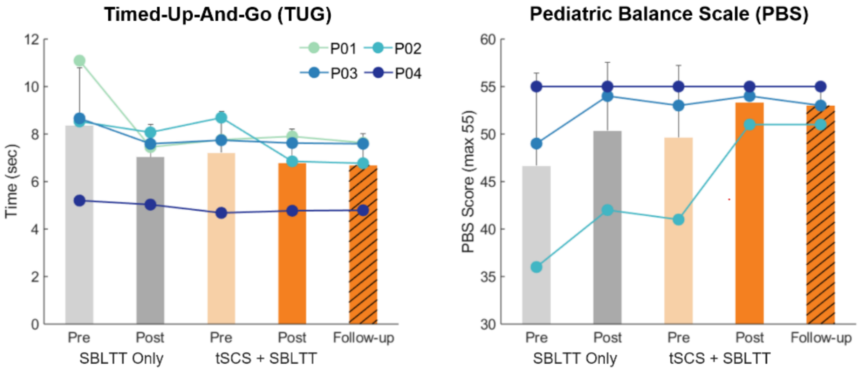


(A) (B)

Supplemental Figure 1. A) Time taken to complete Time-Up-And-Go (TUG) reduced for all participants during SBLTT only and was maintained during tSCS + SBLTT. B) All participants improved on the Pediatric Balance Scale (PBS) during both interventions or reached the maximum score of the assessment. Note the axes limits are from 30-60 points. PBS was not recorded for P01.

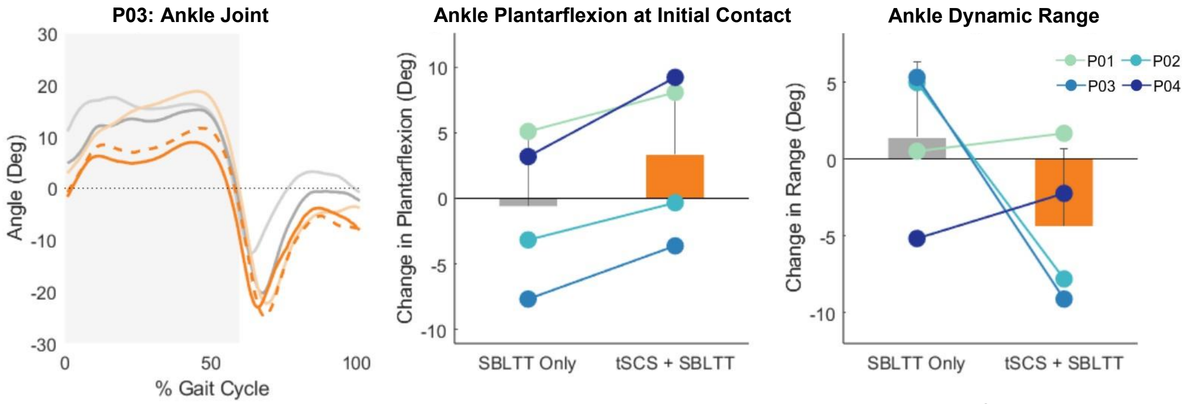


(A) (B) (C)

Supplemental Figure 2. Changes in joint kinematics at the ankle showing A) an example trajectory over the gait cycle from P03, B) change in ankle dorsiflexion at initial contact (0-5% of the gait cycle), and C) the dynamic range of the ankle. All measurements were taken during self-selected barefoot walking.
